# Imaging-based patient inclusion model improves clinical trial performance

**DOI:** 10.1101/2021.01.18.21249895

**Authors:** Michal R. Tomaszewski, Shuxuan Fan, Alberto Garcia, Jin Qi, Youngchul Kim, Robert A. Gatenby, Matthew B. Schabath, William D. Tap, Denise K. Reinke, Rikesh J Makanji, Damon R. Reed, Robert J. Gillies

## Abstract

**Purpose:** Success of new cancer therapies relies strongly on effective selection of the target patient populations. We hypothesize that computational analysis of imaging data can be used for patient enrichment in clinical trials and hence aimed to establish the appropriate framework for this analysis.

**Methods:** This was tested among soft-tissue sarcoma (STS) patients accrued into a randomized clinical trial (SARC021) that evaluated the efficacy of evofosfamide (Evo), a hypoxia activated prodrug, in combination with doxorubicin (Dox). Notably, SARC021 failed to meet its overall survival (OS) objective. We tested whether a radiomic biomarker-driven inclusion/exclusion criterion could have been used to result in a significant treatment benefit of the Evo+Dox combination compared to Dox monotherapy. 164 radiomics features were extracted from 303 SARC021 patients with lung metastases, divided into training and test sets.

**Results:** A single radiomics feature, Short Run Emphasis, was identified as the most informative. Combined into a model along with histological classification and smoking history, an enriched subset (42%) of patients had longer OS in Evo+Dox vs. Dox groups [p=0.01, Hazard Ratio (HR) =0.57 (0.36-0.90)]. Applying the same model and threshold value in an independent test set confirmed the significant survival difference [p=0.002, HR=0.29 (0.13-0.63)], identifying patients most likely to benefit from doxorubicin alone.

**Conclusion:** The study presents a first of its kind clinical-radiomic approach for patient enrichment in clinical trials. We show that, had an appropriate model been used for selective patient inclusion, SARC021 trial could have met its primary survival objective for patients with metastatic STS.

## Introduction

In the last decade, there has been an explosion in the use of advanced image analysis with machine learning, known as “Radiomics” ^1, 2^. Radiomic analyses of cancer can be used to stage, prognose patient outcome, predict response to specific therapies and, most recently, to inform therapeutic choices ^3^ with increasing connectivity between image features and tumor biology ^4^. We hypothesized that radiomic approaches can be used in clinical trials for patient enrichment. We tested this hypothesis in a retrospective analysis of data from the SARC021 ^5^ phase 3 clinical trial in metastatic soft tissue sarcoma that compared overall survival (OS) in cohorts treated with doxorubicin (Dox) to those treated with Dox + Evofosfamide (Evo), a hypoxia activated pro-drug of a brominated version of isophosphoramide mustard (NCT01440088). Although Dox+Evo had shown promise for sarcoma in a phase II study ^6^, the phase 3 trial failed to meet its threshold of increased OS in the Dox+Evo cohort ^5^.

Soft tissue sarcomas are a heterogeneous group of malignancies originating in mesenchymal tissue that most often metastasize to the lungs ^7^. An historical median OS of 12 months for metastatic soft tissue sarcoma patients has steadily improved to 20.4 months on trials, which may be attributed to better patient selection along with better supportive care and additional options in second line and beyond therapies ^8-10^. In the SARC021 trial, the shifting survival with Dox monotherapy led to the study being under powered ^11^. Biomarkers that can exclude a patient cohort that is likely to benefit from standard therapy would be useful to focus trials on those most likely to benefit from an experimental therapeutic approach. In this first of its kind study, we present a novel quantitative imaging framework that can identify patients most and least likely to benefit from trial enrollment. Radiomic feature extraction is combined with a custom statistical analysis method to create a risk score and a threshold for identification of patients predicted to benefit differentially from Dox monotherapy, who could then be excluded from the trial, leveraging the expected benefit of combination treatment in the remaining sub-population. Such radiomics-based biomarkers could be used as companion diagnostics for treatment decision support of approved agents.

## Methods

### Patient populations

This study was approved by the University of South Florida Institutional Review Board. The analysis includes patients who participated in the TH CR-406/SARC021 multicenter clinical trial of Doxorubicin plus Evofosfamide (Dox+Evo) versus Doxorubicin alone (Dox) in locally advanced, unresectable or metastatic soft-tissue sarcoma. Full trial protocol and results were published by Tap et al. ^5^. A total of 640 patients were enrolled. The primary endpoint of the trial was OS. Survival and clinical data were available for 607 patients, and CT images obtained prior to treatment were available for analysis in 588 patients.

### Patient data and CT images

Patient covariates and CT image were obtained from the Sarcoma Alliance for Research through Collaboration (SARC). Patient data and CT images, obtained prior to treatment, was available for 588 patients. The CT images were uploaded into HealthMyne Quantitative Imaging Decision Support (QIDS) software (https://www.healthmyne.com, Madison, WI), where a radiologist with 10 years of experience (S.F.) identified and segmented all visible lesions. 346 patients were found to have at least one lesion in the lung, the most common metastatic site in the considered cohort (followed by liver lesions, identified in 106 patients), as anticipated ^7^. Only lung patients were included in the study to enable comparison of image features between individuals, and hence the use of radiomics. Of these patients, 303 had contrast enhanced CT scans of the lung which could be analyzed, and these were used for quantification. This total cohort of 303 patients used in this study was randomly divided 70:30 into training and test sets using the *sample* function in R version 4.0.2. The test set was sequestered until the final model was developed in the training set for its most stringent validation and increased reproducibility compared to cross-validation approaches ^12^. Robustness of the feature selection to the training/test split was confirmed as described in Supplementary Materials.

### Radiomic feature extraction

Anonymized imaging data and segmentation structures in DICOM format were retrieved from Healthmyne servers. Details of image pre-processing are described in Supplementary Materials. For each patient, a total of 163 features were calculated in 3D using standardized algorithms from the Image Biomarker Standardization Initiative (IBSI) v5 ^13^. The radiomic features included statistical, histogram, shape & size, Grey Level Cooccurrence Matrix (GLCM), Grey Level Run Length Matrix (GLRLM), Grey Level Size Zone Matrix (GLSZM) and Neighboring Grey Tone Difference Matrix (NGTDM) features, as well as 16 peritumoral features as described before ^14^. Laws and Wavelet features were not extracted due to their poor reproducibility reported in previous studies ^15^. As standard in radiomic studies ^16^, to ensure the radiomic signatures provide additional information compared with tumor volume, the features strongly correlated to volume (Pearson |r|>0.8) were excluded from further analysis, while volume itself was included. Spatial stability of the features was assessed, as described in the Supplementary Methods, and unstable features excluded.

### Feature Selection

The goal of this analysis was to identify the radiomic features and patient covariates differentially associated with OS in the two treatment groups, which was the primary endpoint in the original trial ^5^. A new statistical framework was therefore developed. First, univariable Cox proportional hazards regression analysis was used to assess the degree and direction of statistical association of each feature and covariate with post-treatment OS, separately in Dox and Dox+Evo arms. False discovery rate Benjamini-Hochberg ^17^ correction was applied to the p values of radiomic features to account for multiple testing. For each arm, features and covariates were considered promising in either of the two scenarios: (i) They showed significant association (p<0.05) with survival in one treatment arm AND no association (p>0.30)in the other arm, or (ii) they showed potential association (p<0.30) with survival in both groups in opposite directions (HR >1 in one group and <1 in the other).

Correlations between the remaining features were calculated (Pearson’s correlation coefficient for continuous and Chi Square independence test statistics for categorical variables). For significantly correlated (p<0.05) feature groups, features with lowest univariable Cox regression p value in the corresponding treatment group was retained as a representative of the group, and others excluded to avoid redundant information. If these p-values were exactly equal for several features due to the multiplicity correction, the p-values prior to multiplicity correction were compared. Of the remaining features and covariates, the one with lowest p-value ratio in the two treatment groups (low divided by high) was used in model training.

### Final model construction

The two final sets of features and covariates predicted to be most informative of the differential response to Dox or Dox+Evo were used to build the corresponding separate multivariable Cox proportional hazards regression models. Risk scores that are log-transformed relative risks of death were calculated using the *predict*.*coxph* function in R for all patients in the model training cohort and used to determine threshold for patient virtual inclusion and exclusion from the trial. The process of determining the optimum risk score threshold is described in the results section. Risk score values were predicted for all patients in the test cohort from the final multivariable Cox models constructed as above. The threshold values found to result in best separation of the treatment arms as found in the training set were applied to enrich the test cohort, and survival was compared between the treatment arms in the included subset of the test cohort using log-rank test.

For the Dox model, where patients with high risk score values were expected to perform poorly under Dox, and thus be more likely to favor Dox+Evo treatment, a search for the optimal threshold was performed iteratively including sub-cohorts of patients with risk score above 1^st^, 2^nd^,3^rd^ etc to 97^th^ percentile of the total training cohort, evaluating survival difference between the treatment arms in terms of Cox regression p-value and hazard ratio each time. Thus, the entire range of possible thresholds was interrogated, to check if such selection can lead to significant treatment arm separation and identify an optimal threshold value.

## Results

### Patients

Clinical covariates included in the analysis are listed in **Table 1**, with their description included in **Supplementary Table 1**. Presence of lung metastasis was associated with significantly poorer overall survival in the entire cohort of 607 patients (p=0.007, HR=1.34 (1.09-1.65)). This was not the case for patients with liver metastases, which was the second most common metastatic site (p=0.44). Among patients with lung metastases analyzed in this study, no significant survival difference was observed between the two treatment groups (p=0.8), similarly to the entire cohort (p=0.45). Notably, the number of lung metastases in these patients was also not significantly associated with survival (p=0.15).

**Table 1.**
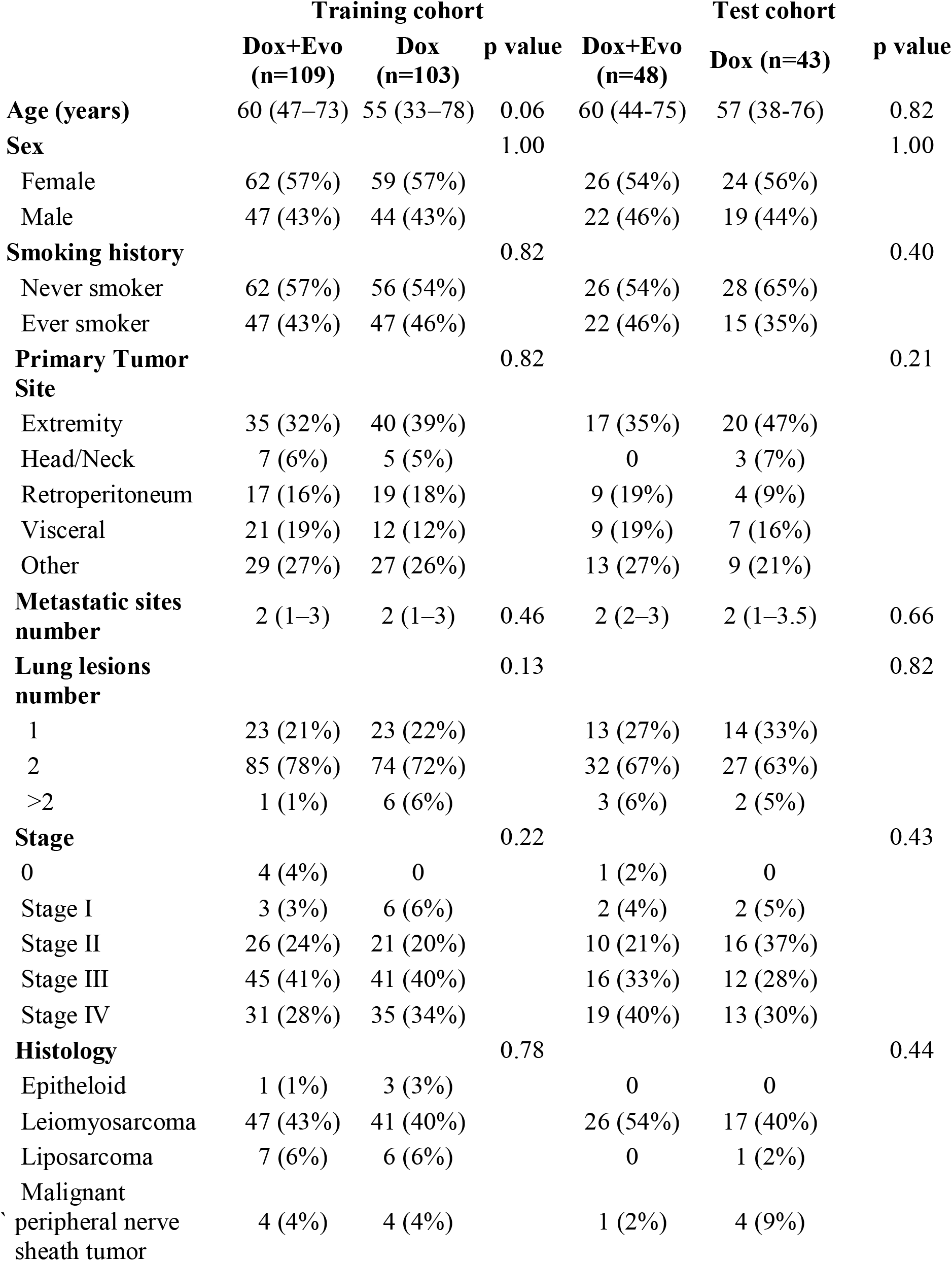
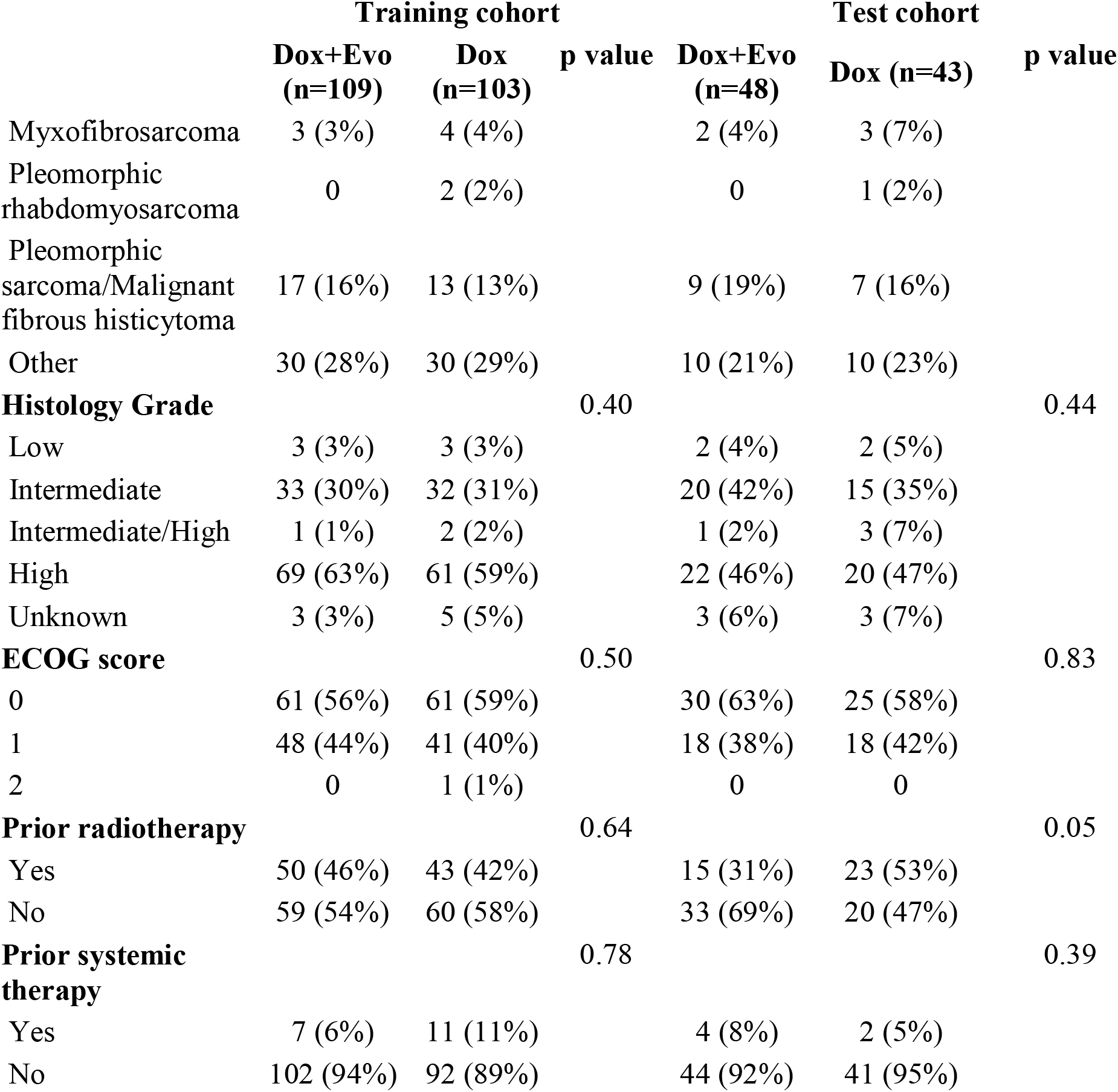
Breakdown of patient characteristics. Numbers are presented for each treatment group in training and test cohort. Data are median (IQR) or n (%). P value by Wilcoxon test (for age) or Chi squared test (all other variables)

|  | Training cohort |  |  | Test cohort |  |  |
| --- | --- | --- | --- | --- | --- | --- |
|  | Dox+Evo<br>(n=109) | Dox<br>(n=103) | p value | Dox+Evo<br>(n=48) | Dox (n=43) | p value |
| **Age (years)** | 60 (47–73) | 55 (33–78) | 0.06 | 60 (44-75) | 57 (38-76) | 0.82 |
| **Sex** |  |  | 1.00 |  |  | 1.00 |
| Female | 62 (57%) | 59 (57%) |  | 26 (54%) | 24 (56%) |  |
| Male | 47 (43%) | 44 (43%) |  | 22 (46%) | 19 (44%) |  |
| **Smoking history** |  |  | 0.82 |  |  | 0.40 |
| Never smoker | 62 (57%) | 56 (54%) |  | 26 (54%) | 28 (65%) |  |
| Ever smoker | 47 (43%) | 47 (46%) |  | 22 (46%) | 15 (35%) |  |
| **Primary Tumor Site** |  |  | 0.82 |  |  | 0.21 |
| Extremity | 35 (32%) | 40 (39%) |  | 17 (35%) | 20 (47%) |  |
| Head/Neck | 7 (6%) | 5 (5%) |  | 0 | 3 (7%) |  |
| Retroperitoneum | 17 (16%) | 19 (18%) |  | 9 (19%) | 4 (9%) |  |
| Visceral | 21 (19%) | 12 (12%) |  | 9 (19%) | 7 (16%) |  |
| Other | 29 (27%) | 27 (26%) |  | 13 (27%) | 9 (21%) |  |
| **Metastatic sites number** | 2 (1–3) | 2 (1–3) | 0.46 | 2 (2–3) | 2 (1–3.5) | 0.66 |
| **Lung lesions number** |  |  | 0.13 |  |  | 0.82 |
| 1 | 23 (21%) | 23 (22%) |  | 13 (27%) | 14 (33%) |  |
| 2 | 85 (78%) | 74 (72%) |  | 32 (67%) | 27 (63%) |  |
| >2 | 1 (1%) | 6 (6%) |  | 3 (6%) | 2 (5%) |  |
| **Stage** |  |  | 0.22 |  |  | 0.43 |
| 0 | 4 (4%) | 0 |  | 1 (2%) | 0 |  |
| Stage I | 3 (3%) | 6 (6%) |  | 2 (4%) | 2 (5%) |  |
| Stage II | 26 (24%) | 21 (20%) |  | 10 (21%) | 16 (37%) |  |
| Stage III | 45 (41%) | 41 (40%) |  | 16 (33%) | 12 (28%) |  |
| Stage IV | 31 (28%) | 35 (34%) |  | 19 (40%) | 13 (30%) |  |
| **Histology** |  |  | 0.78 |  |  | 0.44 |
| Epitheloid | 1 (1%) | 3 (3%) |  | 0 | 0 |  |
| Leiomyosarcoma | 47 (43%) | 41 (40%) |  | 26 (54%) | 17 (40%) |  |
| Liposarcoma | 7 (6%) | 6 (6%) |  | 0 | 1 (2%) |  |
| Malignant peripheral nerve sheath tumor | 4 (4%) | 4 (4%) |  | 1 (2%) | 4 (9%) |  |

|  | Training cohort |  | p value | Test cohort |  | p value |
| --- | --- | --- | --- | --- | --- | --- |
|  | Dox+Evo<br>(n=109) | Dox<br>(n=103) |  | Dox+Evo<br>(n=48) | Dox (n=43) |  |
| Myxofibrosarcoma | 3 (3%) | 4 (4%) |  | 2 (4%) | 3 (7%) |  |
| Pleomorphic<br>rhabdomyosarcoma | 0 | 2 (2%) |  | 0 | 1 (2%) |  |
| Pleomorphic<br>sarcoma/Malignant<br>fibrous histiocytoma | 17 (16%) | 13 (13%) |  | 9 (19%) | 7 (16%) |  |
| Other | 30 (28%) | 30 (29%) |  | 10 (21%) | 10 (23%) |  |
| **Histology Grade** |  |  | 0.40 |  |  | 0.44 |
| Low | 3 (3%) | 3 (3%) |  | 2 (4%) | 2 (5%) |  |
| Intermediate | 33 (30%) | 32 (31%) |  | 20 (42%) | 15 (35%) |  |
| Intermediate/High | 1 (1%) | 2 (2%) |  | 1 (2%) | 3 (7%) |  |
| High | 69 (63%) | 61 (59%) |  | 22 (46%) | 20 (47%) |  |
| Unknown | 3 (3%) | 5 (5%) |  | 3 (6%) | 3 (7%) |  |
| **ECOG score** |  |  | 0.50 |  |  | 0.83 |
| 0 | 61 (56%) | 61 (59%) |  | 30 (63%) | 25 (58%) |  |
| 1 | 48 (44%) | 41 (40%) |  | 18 (38%) | 18 (42%) |  |
| 2 | 0 | 1 (1%) |  | 0 | 0 |  |
| **Prior radiotherapy** |  |  | 0.64 |  |  | 0.05 |
| Yes | 50 (46%) | 43 (42%) |  | 15 (31%) | 23 (53%) |  |
| No | 59 (54%) | 60 (58%) |  | 33 (69%) | 20 (47%) |  |
| **Prior systemic**<br>**therapy** |  |  | 0.78 |  |  | 0.39 |
| Yes | 7 (6%) | 11 (11%) |  | 4 (8%) | 2 (5%) |  |
| No | 102 (94%) | 92 (89%) |  | 44 (92%) | 41 (95%) |  |

No significant difference in OS was observed between the full training and test cohorts (p=0.40, median OS: 17.4 (15.2-20.6) vs. 20.4 (14.0-26.9) training vs test). No significant differences were also seen between Dox and Dox+Evo treatment groups in the training (p=0.67, HR=1.07 (0.78-1.49) median OS: 18.3 (12.6-21.2) vs. 17.2 (15.2-22.1) months Dox vs. Dox+Evo) or test cohorts (p=0.30, HR=1.31 (0.46-1.23) median OS: 23.3 (16.5-31.8) vs. 14.9 (11.1-27.2) months Dox+Evo vs. Dox).

### Feature stability

Concordance coefficients describing the spatial stability of the features were calculated, showing significant heterogeneity between and within feature classes. All results are visualized in **Supplementary Figure 1** and detailed in **Supplementary Table 2**. As expected, shape features remained relatively unchanged, while statistical and histogram features were on average quite strongly affected by choice of ROI. Certain texture features, especially these related to Inverse Difference and Run Length, showed high robustness. Based on this exercise, 54 features with particularly poor robustness (CCC <0.5) were excluded from further analysis. In addition, 12 features strongly correlated with tumor volume (Pearson Correlation Coefficient >0.8) were represented by a single volume feature, leaving 97 features, 81 intratumoral and all 16 peritumoral features.

### Feature selection

Results of univariable Cox proportional hazards regression, performed separately in the Dox and Dox+Evo treatment groups to identify those shortlisted radiomic features and clinical covariates differently associated with OS, are shown in **Table 2**. Among clinical covariates, the histological classification of the primary tumor, tumor grade and prior radiotherapy were significantly associated with survival in the Dox, but not in the Dox+Evo groups. Following elimination of correlated features, histology and prior radiotherapy remained, and histology was chosen to be included in the model due to lower p value ratio (0.015 vs. 0.029). There were no clinical features significantly related to survival in the Dox+Evo group. A history of smoking was significantly associated with shortened survival (p=0.04, HR=1.62 (1.01-2.61)) in the Dox group yet was insignificantly associated with longer survival in the Dox+Evo group (p=0.17, HR=0.73 (0.46-1.15).

**Table 2.** Association with survival in radiomic features and clinical covariates by treatment arm. Univariable Cox regression model was applied separately in the Doxorubicin + Evofosfamide (Dox+Evo) and Doxorubicin only (Dox) treatment arms to calculate the p value (‘p Evo’ and ‘p Dox’ respectively) and hazard ratio (‘HR Evo’ and ‘HR Dox’ together with 95% Confidence Intervals) for the relationship of each feature and covariate with overall survival. P values below 0.05 are highlighted in red, while these above 0.30 are highlighted in green. Hazard Ratios for categorical variables were calculated against most common category. Full table is available in a spreadsheet.

No features or covariates were found to be significant in the Dox+Evo and not in the Dox group. Three uncorrelated radiomic features were found significantly associated with survival in the Dox but not in the Dox+Evo group: Short Run Emphasis, Normalized Run Length Nonuniformity and Small Zone Emphasis. Of these features, Short Run Emphasis, a measure of heterogeneity, showed the lowest ratio of p value in the Dox to the p value in Dox+Evo groups (p=0.018 and p=0.63 respectively), and was chosen for training a prediction model of post-treatment survival.

### Multivariable model

The three features identified above (histology, non-smoking history, and radiomic Short Run Emphasis) were combined in a multivariable Cox model trained on the Dox cohort of the training data set, producing a highly significant signature (p=0.0001) of survival. Details of the model are shown in **Supplementary Table 3**. No significant correlation between residuals and time was measured (p=0.46, 0.45, 0.71 and 0.58 for SRE, histology, smoking history, and global test respectively) validating the proportional hazards assumption for Cox model use. No corresponding model was developed in the Dox+Evo group, as no clinical or radiomic features specific to this treatment arm were identified. The Dox model was used to predict risk scores for the entire training set cohort, including patients in the Dox+Evo group, providing a predicted measure of risk of death if the Dox treatment was applied to all patients. Patients with highest risk scores for Dox monotherapy (i.e. worse outcome) are expected to benefit the most from the alternative (Dox+Evo) treatment, and hence they should be included in the trial. Conversely, patients with a low Dox risk score should be excluded and undergo Dox monotherapy instead. Such a patient enrichment strategy for the trial would thus be expected to result in an improved treatment benefit of Dox+Evo in the included patients. To assess this, we performed the log-rank test for difference in survival between Dox vs. Dox+Evo as a function of the risk score threshold for the remaining patients whose score was above that threshold. The schematic of the process is shown in **Figure 1**. As described in Methods, the threshold separating high-from low-risk groups was incrementally increased to identify an optimum that reached a significant (p<0.05) difference in OS, while including the largest fraction of patients. The results of this analysis are shown in **Figure 2**, demonstrating that increasingly different OS can be observed for the two treatment groups when patients with low-risk scores are excluded from the analysis (**Figure 2A**).The smaller p-values encountered with increasing thresholds were consistent with decreasing HR <1 (**Figure 2B**), showing increasingly significant treatment benefit of Dox+Evo vs. Dox with more stringent inclusion criteria. A threshold of 1.00 allowed inclusion of 42% of the initial training cohort in the trial and showed a significant advantage of Dox+Evo over Dox [p=0.017, HR=0.57 (0.36-0.90)]. This result was visualized in divergent Kaplan Meier curves for the treatment groups in the included patients and longer survival in the Dox+Evo group (**Figure 2C**, median survival 15.8 (14.2-21.4) vs 9.1 (7.6-13.7) months Dox+Evo vs Dox) and a reverse trend for the excluded patients (**Figure 2D**, median survival 20.9 (14.7-27.2) vs 27.0 (20.1-Not estimable(N.E)) Dox+Evo vs Dox, p=0.036), with Dox treated group showing significantly longer survival, possibly related to the lack of Evo response in the subgroup and its added toxicity ^5^. No difference was observed in the whole training cohort (**Figure 2E**). These figures make apparent that the most significant difference (p<10^−5^) between included and excluded groups is their response to Dox monotherapy. Indeed, the difference in survival between included and excluded groups in the Dox+Evo trial arms was insignificant (p=0.48). A model using only clinical features as input was also trained and its performance evaluated as above. At the same inclusion rate as the radiomic-clinical model (42%), this approach does not show a significant survival difference between treatment arms (p=0.07, HR=0.66 (0.42-1.03)), only reaching significance at a much lower proportion of included patients (32%, p=0.0007, HR= 0.39 (0.23-0.67) (see **Supplementary Figure 2** for performance details). Although the Short Run Emphasis feature by itself did not significantly discriminate (minimum p-value 0.097 including only patients with SRE<0.84, **Supplementary Figure 3**), it added to the significance of clinical features and thus increased the number of potential patients on trial from 32% to 42%. Selection based on lesion volume, routinely used in radiological analysis, could not separate patients likely to respond significantly differently to Dox and Dox+Evo treatments, neither through application of upper nor lower volume threshold (see **Supplementary Figure 4**).

**Figure 1.**
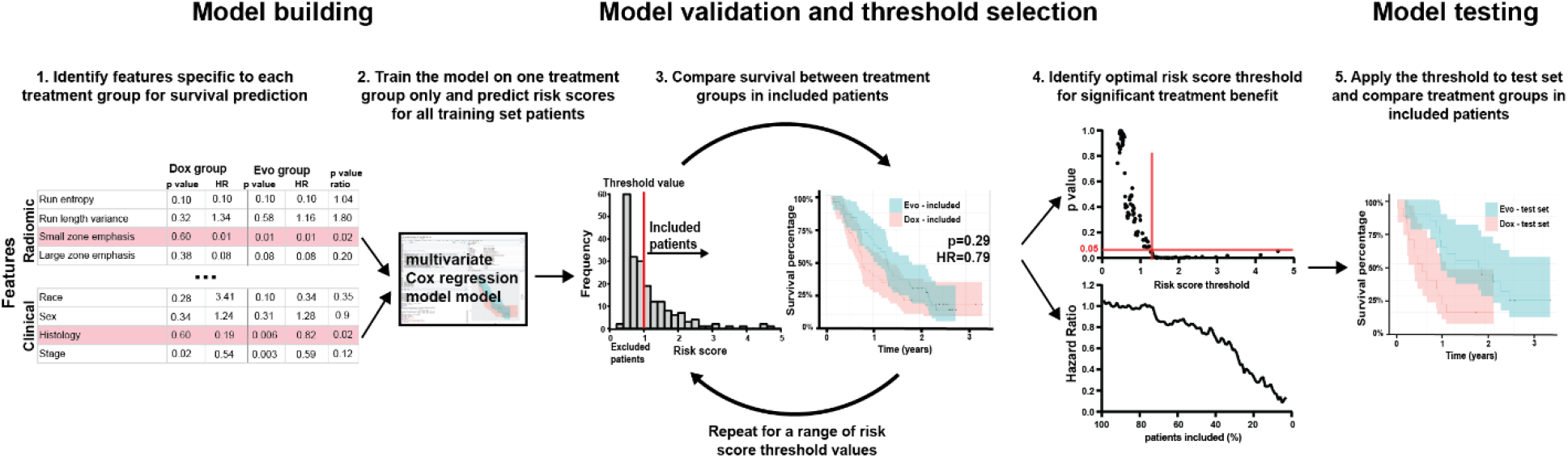
Patient inclusion model. Patient selection into the trial based on Dox group survival was executed according to the following method: firstly (1) radiomic and clinical features associated in training cohort with survival in Dox but not Dox+Evo treatment group were included in a multivariable Cox regression model (2), trained on Dox treated patients. The risk score assigned by the model to each training set patient was then used as a biomarker for inclusion into analysis, iteratively calculating the p value and hazard ratio for survival comparison between treatment arms depending on minimum risk score threshold (3). If available, threshold corresponding to significant (p value<0.05) treatment benefit of Dox+Evo at highest percentage of patients included was chosen (4), and subsequently tested in the test cohort (5), with risk scores assigned by the multivariable Cox model developed in step (2). A corresponding model can also be developed based on Dox+Evo group survival, using a maximum risk score threshold.

**Figure 2.**
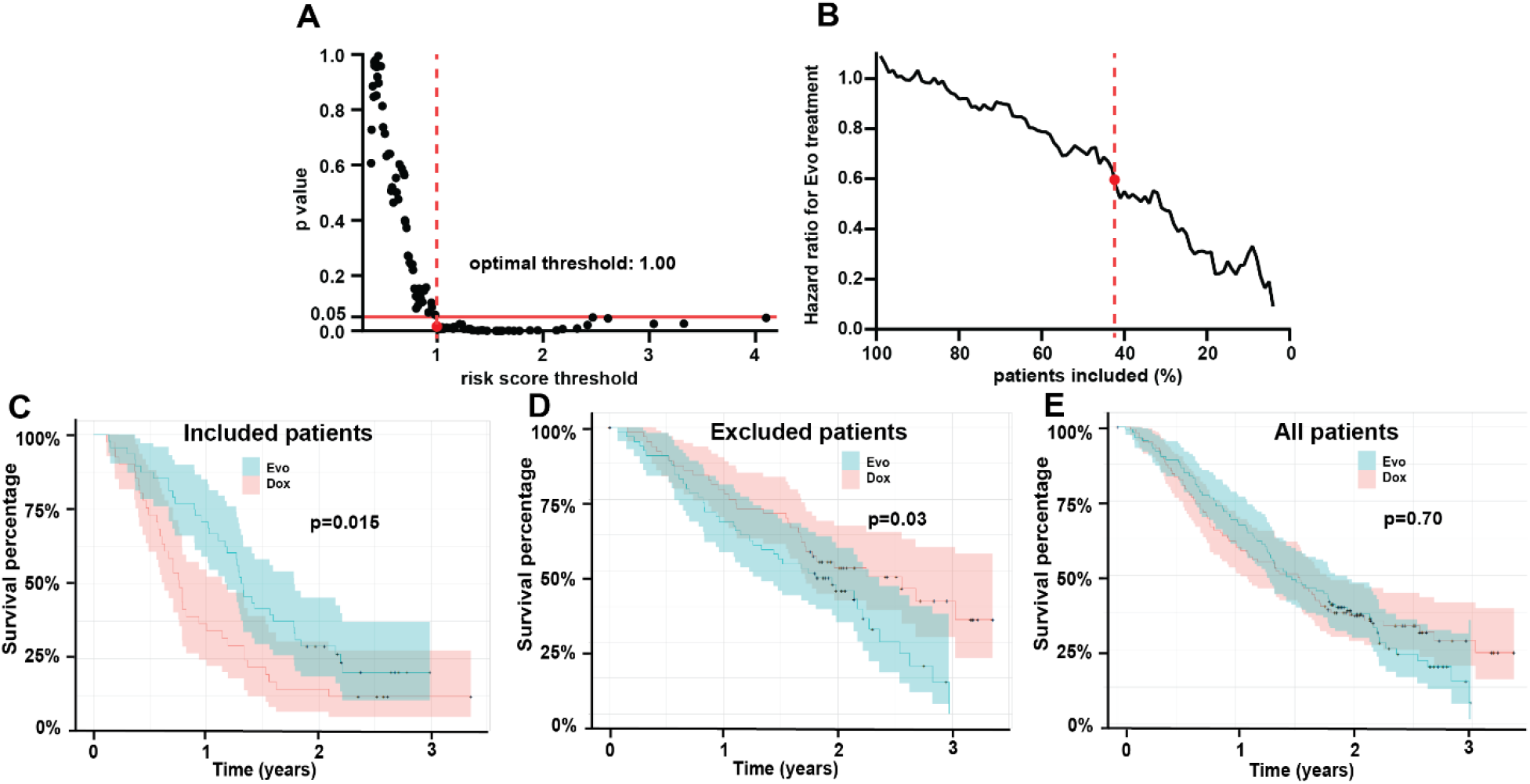
Multivariable Cox model enables selection of patients who benefit from Evofosfamide+Doxorubicin in training cohort. Quantification of the p value of overall survival difference in the training cohort between the Evofosfamide+Doxorubicin (Dox+Evo) vs. Doxorubicin alone (Dox) treatment arms depending on the minimum risk score for patient inclusion, as predicted by the model (A), shows a risk score threshold of 1.00 at which Doxorubicin +Evofosfamide (Dox+Evo) group shows significantly longer OS (p<0.05). Exclusion of patients with high risk scores leads to monotonic decrease in the hazard ratio (B), and the 1.00 risk score threshold corresponds to inclusion of 42% of patients in the trial (indicated by red dotted line). The Kaplan-Meier plots by treatment arms show significantly better OS in the included (C) and significantly worse OS in the (D) excluded patients for the Dox+Evo treatment compared to Dox only. In all training set patients (E) no difference between the arms was observed.

### Model testing

The multivariable Cox model trained in the above section was used to predict risk scores for all patients in the test cohort. Similar to the training cohort, an increase in minimum risk score threshold for inclusion lead to a monotonic decrease in p value (**Figure 3A**) and HR (**Figure 3B**) for the overall survival comparison between treatment groups. Applying the threshold of 1.00 determined *a priori* in the training set as the optimum threshold for inclusion, showed a significantly better survival in the Dox+Evo vs the Dox treated group [p=0.002, HR=0.29 (0.13-0.63) **Figure 3C**]. As in the training cohort, this was associated with an increased median survival of 22.8 (12.3-N.E) for Dox+Evo vs. 6.3 (3.1-13.7) for Dox. As shown in **Figure 3D** the differences in the two treatment arms for the remaining excluded patients was insignificant for both OS (p=0.72) and median survival 26.0 (18.4-N.E) vs. 27.2 (18.6-N.E), similar to the starting whole test cohort (**Figure 3E**). The threshold corresponded to inclusion of 38% of the test cohort. As in the training set (**Figure 2C**,**D**), the selection by risk score threshold separated patients who did and did not respond to Dox (p<10^−4^), whereas it did not discriminate (p=0.54) responses of the Dox+Evo group. The plot of p-value *vs*. inclusion threshold (**Figure 3A**) shows a matching profile of improving treatment benefit of the Dox+Evo treatment (because of decreasing effectiveness of Dox) with increasing risk score, further supporting the model and the use of radiomics in patient selection. The similar proportions of ‘included’ patients in the training and test set (42% and 39% respectively) support the validity of the model and its statistical consistency between the sets ^1^.

Repeated random draw of 70% of all analyzed patients confirmed the robustness of associations between the final model variables and survival to training/test split, as described in Supplementary Materials.

**Figure 3.**
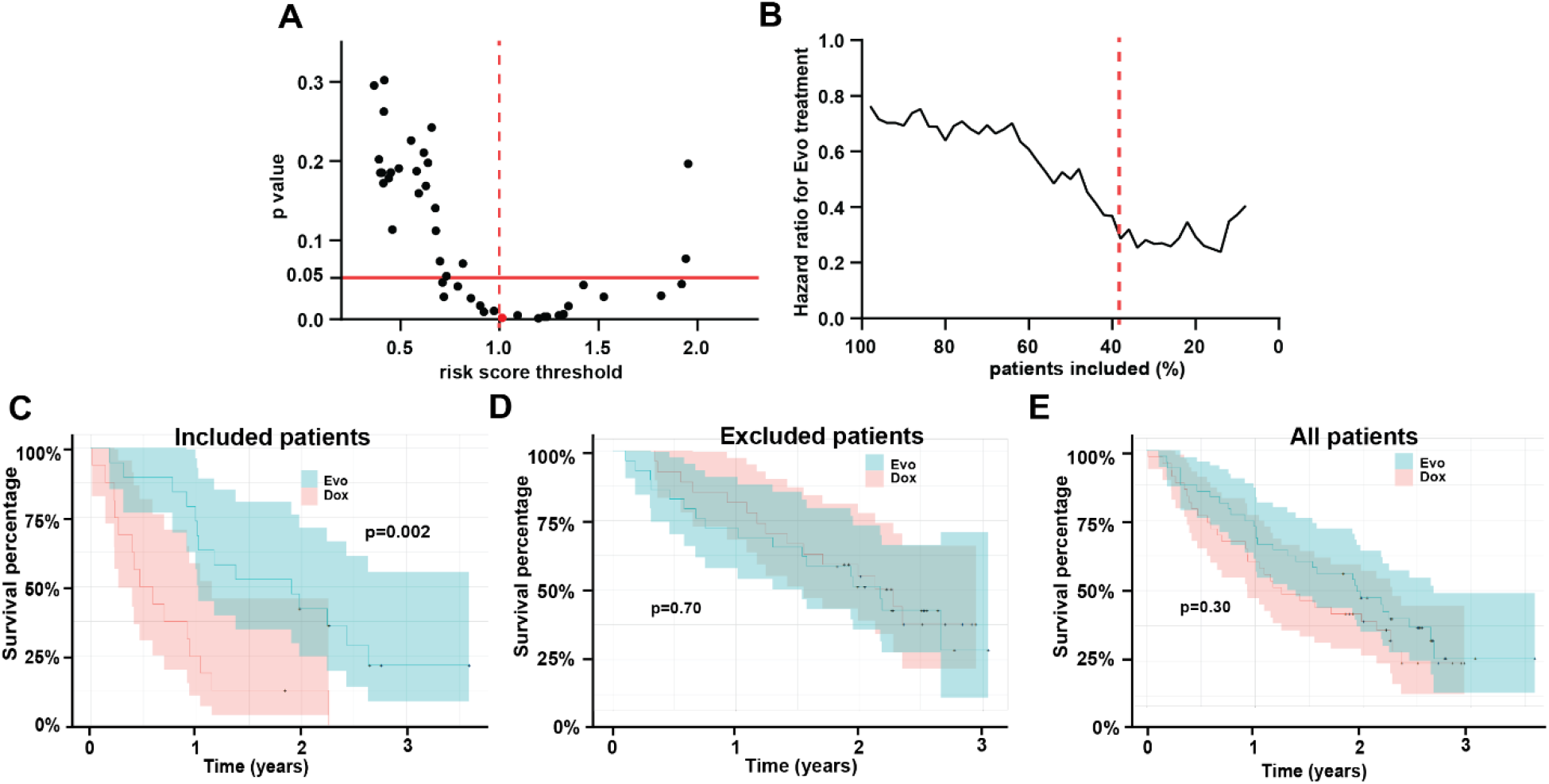
Results in the test cohort confirm the validity of the model. Risk scores predictions in the test cohort based on a multivariable Cox model trained on Dox treated training cohort patients can be used to identify patients who would benefit from Dox+Evo treatment. Graph in (A) shows that increasing the minimum risk score of patients included in the analysis leads to a stronger difference in survival between the treatment groups, as described by the p value of the comparison. For the risk score threshold of 1.00, a highly significant difference is observed (red point and dotted line), which corresponds to a decreased hazard ratio of the combination vs standard therapy (B). These differences are apparent from the Kaplan-Meier curve in the included patients (C) showing significantly longer survival in the Dox+Evo group, while the excluded patients (D), or all test set patients (E) show no difference in survival between treatment groups.

### Model interpretation

The hazard ratios for the constituent variables in the final model, as shown in **Supplementary Table 3**, can be used to shed light on the underlying relationships. Here HR>1 suggests a poor prognostic factor for Dox monotherapy, with its enrichment improving the potential treatment benefit of Evo addition. Most histologies, except for a relatively rare Myxofibrosarcoma, show high HR vs. Leiomyosarcoma, an observation in line with the lack of response to Evosfosfamide in Leiomyosarcoma noted in the original SARC021 trial ^5^. Excluding this common histology from the trial does not result in a significant OS benefit in the cohort of the remaining patients, both in the training (p=0.22, HR=0.77 (0.51-1.16)) or full dataset (p=0.39, HR=0.86 (0.61-1.21)). A past history of smoking is a trending poor prognostic factor in the model, and given its lack of relevance in the Dox+Evo arm, including only current or ex-smokers in the analysis would result in an improved benefit of Dox+Evo in the training cohort (p=0.12, HR=0.68 (0.42-1.10)). Interestingly, conversely the never-smokers of the cohort show significantly better survival on standard Dox compared to Dox+Evo (p=0.049, HR=1.52 (1-2.47)). Analysis of the full cohort of trial patients confirmed the relevance of smoking history in the treatment response. For patients without lung metastases, not included in this study, Dox treatment showed no benefit for never-smokers (p=0.89, HR=0.97 (0.59-1.57)) while a trending benefit of Dox+Evo was observed in Ex/Current smokers (p=0.11, HR=1.53 (0.92-2.54)).

Given the complexity of the question, directly interpreting the imaging information in the model may be challenging. The analysis of p-value *vs*. the value threshold (as shown in **Figures 2** and **3** for risk scores) for the Short Run Emphasis (SRE) feature, showed that the treatment benefit of Dox+Evo is maximized if only patients with tumors of low SRE are included (minimum p= 0.097, HR=0.62 (0.35-1.09) at SRE<0.845, **Supplementary Figure 3**). Taking the entire dataset into account, the p value at this threshold reaches p=0.01, reinforcing the relevance of this radiomic feature in patient selection. The multivariable model developed above also favors low SRE values, as shown in **Figure 4A** for both training and test cohorts. The biological meaning of the SRE feature is not obvious, but inferences can be made. For example, comparing representative tumors with extreme SRE values reveals visual differences. In line with the model, a patient censored after over 2.5 years showed a very low SRE in the lung lesion (**Figure 4B**) at baseline; and this is visually associated with regularity and homogeneity of the mass. Conversely, another patient deceased on Dox therapy less than 5 months after enrollment presented a lung lesion with high SRE and highly heterogeneous appearance (**Figure 4C**). While these show extremes, the value of using a quantitative SRE threshold is to identify those patients whose scans may be less obvious and hence, equivocal. SRE was not shown to correlate with the CT image characteristics, with Pearson correlation coefficient to in-plane voxel size r= −0.03, and Wilcoxon test p value=0.70 between the scans of slice thickness ≤3 and >3, supporting the biological origin of the signal. The feature also showed particularly high spatial stability (Concordance coeff. 0.90, 80^th^ percentile).

**Figure 4.**
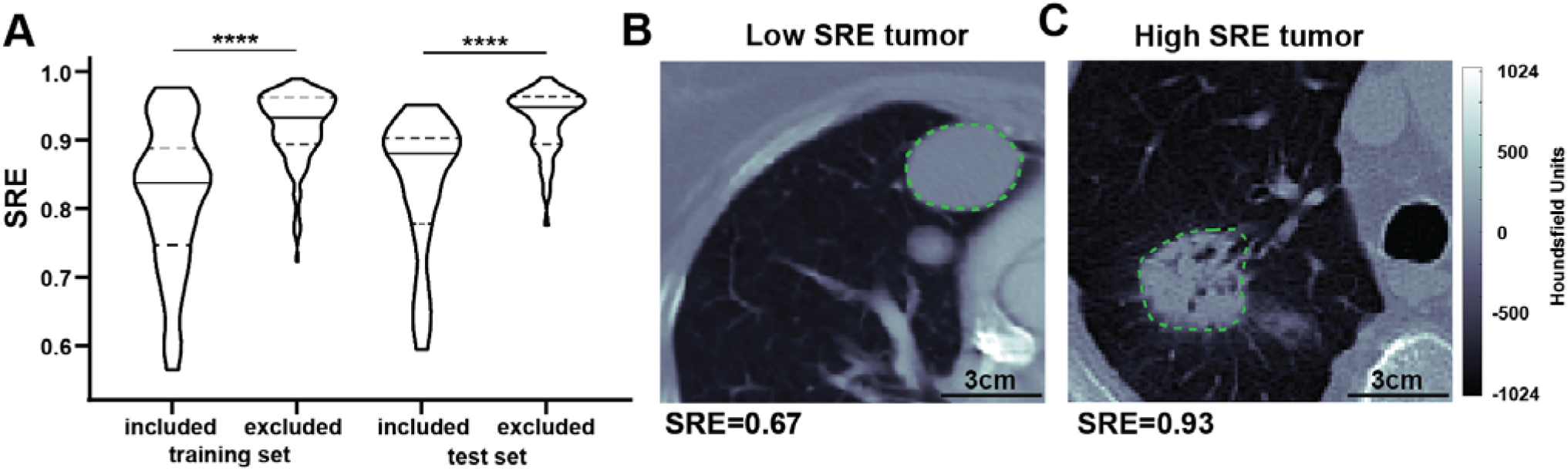
Differences in radiomic features can be apparent visually. The model for selection of patients likely to benefit from Evofosfamide treatment favored low Short Run Emphasis (SRE) radiomic feature for proposed inclusion into the trial. As shown in the violin plot (A), significantly lower SRE is observed in the included vs. excluded patient groups both in training and test cohorts. Qualitatively, a representative tumor with low Short Run Emphasis (SRE, B) appears more regular and homogeneous in a contrast enhanced CT scan than a corresponding tumor with similar volume (15.0 vs 16.5ml respectively), and relatively high SRE (C), which shows higher intratumor heterogeneity. In the violin plot a solid line indicates median while dotted lines indicate 25^th^ and 75^th^ percentile. **** p<0.0001

## Discussion

Herein, we identified a novel radiomic model, based on the combination of pre-treatment CT data and clinical information, that predicts patients that would have relatively long OS with Dox monotherapy. The strong predictive model of Dox monotherapy response shows significant promise for both clinical care and more optimal patient selection in future trials. The sarcoma community has long sought additional efficacious agents in metastatic soft tissue sarcomas with large trials dedicated to alkylators such as ifosfamide ^18^, and derivatives like palifosfamide ^19^ that have shown increased response rates but not overall survival benefit. Additionally, albeit in localized soft tissue sarcomas, a recent trial comparing histology directed therapy compared to doxorubicin concluded that doxorubicin and ifosfamide remain the standard first line agent for all tested histologies ^20^.

In the SARC021 trial this enrichment for patients unlikely to benefit from doxorubicin would have improved the relative effect of evofosfamide. These results were successfully validated in the test set and, if applied, the phase 3 trial would have been successful. The failure of the SARC21 trial is at least in part due to a shifting OS for Dox monotherapy that is likely multifactorial; inclusive of better patient selection, understanding of histologies and patients most likely to benefit from systemic therapy, improved supportive care and additionally available subsequent therapies ^5, 18, 19^.

Patient selection for drug trials remains a challenge in clinical trial design. In the study, it is notable that the inclusion/exclusion strategy was generated from readily available standard-of-care images and clinical data. Herein, radiomic methods ^16^ in combination with novel statistical analysis was used to provide and validate a patient inclusion framework based on widely available standard of care imaging data in a retrospective cohort. While radiomics methods have been used to predict patient survival following different treatments ^21-24^, and correlate to tumor hypoxia ^25, 26^, this is the first study to derive the prognostic radiomic features and multivariable models required to discriminate between two therapeutic arms and determine the optimal population to most benefit from the novel Evo intervention.

The analysis in this study focused on patients with lung metastases, as they are the most common and deadly metastatic site. This study highlighted the value of combining tumor histology, patient smoking history, and CT imaging information for trial population enrichment. Notably neither clinical nor imaging information alone were sufficient to show significant benefit of Evo+Dox in the selected cohort, emphasizing the value of the quantitative model framework proposed in this study, and specific identification of the population of interest. Interestingly, current or ex-smokers were more likely to benefit from the addition of Evofosfamide than those who never smoked. This observation is consistent with the hypoxia-targeting action of the drug, as smoking is known to exacerbate tumor hypoxia through reduction of blood oxygen carrying capacity ^27^, especially in the lungs, which may lead to improved response to hypoxia targeted treatment in these tumors compared to standard therapy, contributing to the final proposed model. Notably, while there were a number of prognostic features associated with positive outcomes in both groups, there were no features were associated with survival in the Dox+Evo cohort independent of the Dox group. This suggests that the biological factors that govern Evo response may not be related sufficiently strongly to the information available from clinical and imaging data.

Significant strength of the developed model comes from the heterogeneity of the training and testing data. Obtained in a multicenter international trial, the CT imaging was performed on multiple systems with varying acquisition parameters, making the final signature more robust and generalizable. However, there are some limitations to the presented study. As radiomic patterns and relationships with outcome are known to vary significantly between disease sites, the image quantification in this study was confined to the metastatic lesions in the lung. Lung metastases are both the most common site and leading cause of death in sarcoma ^7^. The advantage of this analysis is that there was not special radiologic protocol needed for image acquisition and indeed no planning for this radiomic analysis was considered in the trial design. The dataset was collected as part of the trial and the hypothesis and methods applied to this dataset. While the original lack of survival benefit of the novel treatment was conserved in this sub-cohort, in the future the analytical approach proposed should be extended to include other metastatic sites. An observational trial in soft tissue sarcoma patients bearing lung metastases treated with Dox is planned at our institution to validate the model and understand its biological underpinnings ^4^, prospectively comparing the model-calculated risk score to overall survival. The prospective data can be used to support this radiomic biomarker as a companion diagnostic or integrated biomarker for patient selection.

In summary, in this work we demonstrate for the first time that machine learning can be used to predict differential survival to distinct treatment regimens. We show that radiomic analysis of CT imaging data can be used in combination with clinical information to develop a first of its kind model capable of identifying soft tissue sarcoma patients likely to benefit from novel combination of Doxorubicin+Evofosfamide *vs*. standard Doxorubicin. Application of the proposed model shows that should patient selection be performed a significant survival benefit could have been observed in an otherwise negative Phase 3 trial. Used prospectively, this approach may in the future improve the chance of determining efficacy of novel therapeutic regimens through better patient selection and guide therapeutic decisions for all metastatic STS through actionable, personalized, image-based, survival prediction.

## Data Availability

The analysis code and processed anonymized data may be available on request.

## Acknowledgements

The authors would like to thank Dr Benjamin Haibe-Kains and Ms Caryn Geady at the University of Toronto for helpful feedback on the manuscript.

## References

1. Lambin P, Leijenaar RTH, Deist TM, et al. Radiomics: the bridge between medical imaging and personalized medicine. Nat Rev Clin Oncol. Dec 2017;14(12):749–762. doi:10.1038/nrclinonc.2017.14110.1038/nrclinonc.2017.141. Epub 2017 Oct 4.

2. Gillies RJ, Kinahan PE, Hricak H. Radiomics: Images Are More than Pictures, They Are Data. Radiology. Feb 2016;278(2):563–77. doi:10.1148/radiol.201515116910.1148/radiol.2015151169. Epub 2015 Nov 18.

3. Mu W, Jiang L, Zhang J, et al. Non-invasive decision support for NSCLC treatment using PET/CT radiomics. Nat Commun. Oct 16 2020;11(1):5228. doi:10.1038/s41467-020-19116-x

4. Tomaszewski MR, Gillies RJ. The biological meaning of radiomic features. Radiology. 2020;in press

5. Tap WD, Papai Z, Van Tine BA, et al. Doxorubicin plus evofosfamide versus doxorubicin alone in locally advanced, unresectable or metastatic soft-tissue sarcoma (TH CR-406/SARC021): an international, multicentre, open-label, randomised phase 3 trial. The Lancet Oncology. 2017 Aug 2017;18(8)doi:10.1016/S1470-2045(17)30381-9

6. Chawla SP, Cranmer LD, Van Tine BA, et al. Phase II study of the safety and antitumor activity of the hypoxia-activated prodrug TH-302 in combination with doxorubicin in patients with advanced soft tissue sarcoma. Journal of clinical oncology : official journal of the American Society of Clinical Oncology. 10/10/2014 2014;32(29)doi:10.1200/JCO.2013.54.3660

7. Billingsley KG, Burt ME, Jara E, et al. Pulmonary metastases from soft tissue sarcoma: analysis of patterns of diseases and postmetastasis survival. Annals of surgery. 1999 May 1999;229(5)doi:10.1097/00000658-199905000-00002

8. Tap WD, Wagner AJ, Schoffski P, et al. Effect of Doxorubicin Plus Olaratumab vs Doxorubicin Plus Placebo on Survival in Patients With Advanced Soft Tissue Sarcomas: The ANNOUNCE Randomized Clinical Trial. JAMA. Apr 7 2020;323(13):1266–1276. doi:10.1001/jama.2020.1707

9. Seddon B, Strauss SJ, Whelan J, et al. Gemcitabine and docetaxel versus doxorubicin as first-line treatment in previously untreated advanced unresectable or metastatic soft-tissue sarcomas (GeDDiS): a randomised controlled phase 3 trial. Lancet Oncol. Oct 2017;18(10):1397–1410. doi:10.1016/S1470-2045(17)30622-8

10. Lorigan P, Verweij J, Papai Z, et al. Phase III trial of two investigational schedules of ifosfamide compared with standard-dose doxorubicin in advanced or metastatic soft tissue sarcoma: a European Organisation for Research and Treatment of Cancer Soft Tissue and Bone Sarcoma Group Study. Journal of clinical oncology : official journal of the American Society of Clinical Oncology. 07/20/2007 2007;25(21)doi:10.1200/JCO.2006.09.7717

11. Lindner LH. Hypoxia-activated prodrug: an appealing preclinical concept yet lost in clinical translation. The Lancet Oncology. 2017 Aug 2017;18(8)doi:10.1016/S1470-2045(17)30401-1

12. Papanikolaou N, Matos C, Koh DM. How to develop a meaningful radiomic signature for clinical use in oncologic patients. Cancer imaging : the official publication of the International Cancer Imaging Society. 05/01/2020 2020;20(1)doi:10.1186/s40644-020-00311-4

13. Zwanenburg A, Leger S, Vallières M, Löck S. Image biomarker standardisation initiative. 2016/12/21 2016;

14. Tunali I, Stringfield O, Guvenis A, et al. Radial gradient and radial deviation radiomic features from pre-surgical CT scans are associated with survival among lung adenocarcinoma patients. Oncotarget. Nov 10 2017;8(56):96013–96026. doi:10.18632/oncotarget.2162910.18632/oncotarget.21629. eCollection 2017 Nov 10.

15. Tunali I, Hall LO, Napel S, et al. Stability and reproducibility of computed tomography radiomic features extracted from peritumoral regions of lung cancer lesions. Med Phys. Nov 2019;46(11):5075–5085. doi:10.1002/mp.1380810.1002/mp.13808. Epub 2019 Sep 23.

16. Yip SS, Aerts HJ. Applications and limitations of radiomics. Physics in medicine and biology. 07/07/2016 2016;61(13)doi:10.1088/0031-9155/61/13/R150

17. Benjamini Y, Hochberg Y. Controlling the False Discovery Rate: A Practical and Powerful Approach to Multiple Testing. Journal of the Royal Statistical Society Series B (Methodological). 1995;57(1):289–300.

18. Judson I, Verweij J, Gelderblom H, et al. Doxorubicin alone versus intensified doxorubicin plus ifosfamide for first-line treatment of advanced or metastatic soft-tissue sarcoma: a randomised controlled phase 3 trial. The Lancet Oncology. 2014 Apr 2014;15(4)doi:10.1016/S1470-2045(14)70063-419.

19. Ryan CW, Merimsky O, Agulnik M, et al. PICASSO III: A Phase III, Placebo-Controlled Study of Doxorubicin With or Without Palifosfamide in Patients With Metastatic Soft Tissue Sarcoma. Journal of clinical oncology : official journal of the American Society of Clinical Oncology. 11/10/2016 2016;34(32)doi:10.1200/JCO.2016.67.6684

20. Gronchi A, Palmerini E, Quagliuolo V, et al. Neoadjuvant Chemotherapy in High-Risk Soft Tissue Sarcomas: Final Results of a Randomized Trial From Italian (ISG), Spanish (GEIS), French (FSG), and Polish (PSG) Sarcoma Groups. Journal of clinical oncology : official journal of the American Society of Clinical Oncology. 07/01/2020 2020;38(19)doi:10.1200/JCO.19.03289

21. Chetan MR, Gleeson FV. Radiomics in predicting treatment response in non-small-cell lung cancer: current status, challenges and future perspectives. European radiology. 08/18/2020 2020;doi:10.1007/s00330-020-07141-9

22. Agrawal V, Coroller TP, Hou Y, et al. Radiologic-pathologic correlation of response to chemoradiation in resectable locally advanced NSCLC. Lung cancer (Amsterdam, Netherlands). 2016 Dec 2016;102 doi:10.1016/j.lungcan.2016.10.002

23. Horvat N, Veeraraghavan H, Khan M, et al. MR Imaging of Rectal Cancer: Radiomics Analysis to Assess Treatment Response after Neoadjuvant Therapy. Radiology. 2018 Jun 2018;287(3)doi:10.1148/radiol.2018172300

24. Kickingereder P, Götz M, Muschelli J, et al. Large-scale Radiomic Profiling of Recurrent Glioblastoma Identifies an Imaging Predictor for Stratifying Anti-Angiogenic Treatment Response. Clinical cancer research : an official journal of the American Association for Cancer Research. 12/01/2016 2016;22(23)doi:10.1158/1078-0432.CCR-16-0702

25. Ganeshan B, Goh V, Mandeville HC, Ng QS, Hoskin PJ, Miles KA. Non-small cell lung cancer: histopathologic correlates for texture parameters at CT. Radiology. Jan 2013;266(1):326–36. doi:10.1148/radiol.12112428

26. Rooney M, Govindan R. The era of big trials is over. The Lancet Oncology. 2013 Jan 2013;14(1)doi:10.1016/S1470-2045(12)70512-0

27. Salem A, Asselin MC, Reymen B, et al. Targeting Hypoxia to Improve Non-Small Cell Lung Cancer Outcome. Journal of the National Cancer Institute. 01/01/2018 2018;110(1)doi:10.1093/jnci/djx160

