## Supplementary Materials for "Imaging-based patient inclusion model improves clinical trial performance"

**Supplementary Methods**

*Image processing*

Anonymized imaging data and segmentation structures in DICOM format were retrieved from Healthmyne servers. Preprocessing was performed using custom scripts in MATLAB 2020a (The Mathworks Inc., Natick, Massachusetts) to convert the files into image matrices and corresponding region of interest (ROI) binary masks. For each patient, the lung lesion with largest ROI volume was chosen for quantification. Image quantification was performed in MATLAB using an in-house extraction toolbox created in MATLAB and C++. Hounsfield units (HU) in all CT images were then resampled into fixed bin sizes of 25 HUs discretized from –1000 to 1000 HU. Due to the multicenter nature of the trial, the images were acquired at a range of voxel sizes, with slice thickness of mainly 3 or 5mm (median: 5mm, IQR:4-6mm), pixel length ranging from 0.5mm to 1.17mm (mean 0.79mm, SD 0.11mm). To account for these differences, the CT images were resampled to a single voxel spacing of 1mm x 1mm x 1mm using cubic interpolation to standardize spacing.

*Feature stability*

Manual lesion segmentation introduces inherent variability into image quantification. While strict rules govern how the outlines should be drawn, some radiomic features are more susceptible to small changes in ROI shape and size than others. These spatially unstable features should be excluded from further analysis to ensure robustness. Feature values were calculated in the tumor ROI as segmented, as well as after its erosion or dilation by 1mm, simulating intra-observer differences in segmentation. Concordance coefficients between the original, shrunk and dilated ROIs were calculated, showing significant heterogeneity between and within feature classes. All results are presented in **Supplementary** **Table 2** and visualized in **Supplementary Figure 3**. As expected, shape features remained relatively unchanged, while statistical and histogram features were on average quite strongly affected by choice of ROI. Certain texture features, especially these related to Inverse Difference and Run Length, showed high robustness. Based on this exercise, 54 features with particularly poor robustness (CCC <0.5) were excluded from further analysis. In addition, 12 features strongly correlated with tumor volume (PCC >0.8) were represented by a single volume feature, leaving 81 intratumoral and all 16 peritumoral features.

*Robustness of feature selection to training/test split*

Robustness of the feature selection to the training/test split was assessed by comparing p-values after 100 more random 70/30 patient selection steps, using the *sample* function in R. This was performed after final model training and testing not to reveal information about the initial independent test set.

This analysis confirmed the model selection from the first split. Short Run Emphasis radiomic feature, as used in the final model, showed significant (p<0.05) association to overall survival in Dox arm in 96/100 cases and no association (p>0.30) in Dox+Evo arm in 96 cases – the most robust of all radiomic features. Out of clinical variables, tumor histology showed no association to survival in Dox+Evo arm in 30/100 splits, and significant association to overall survival in Dox arm in 96/100 splits, the highest number apart from ECOG score which showed the same significant association with survival 100/100 in both Dox and Dox+Evo, and was therefore excluded. As in the final model, Smoking history showed trending opposite association with survival in the Dox and Dox+Evo arms, highlighted in 33/100 splits, not seen in any other radiomic or clinical variables.

**Supplementary Figures**

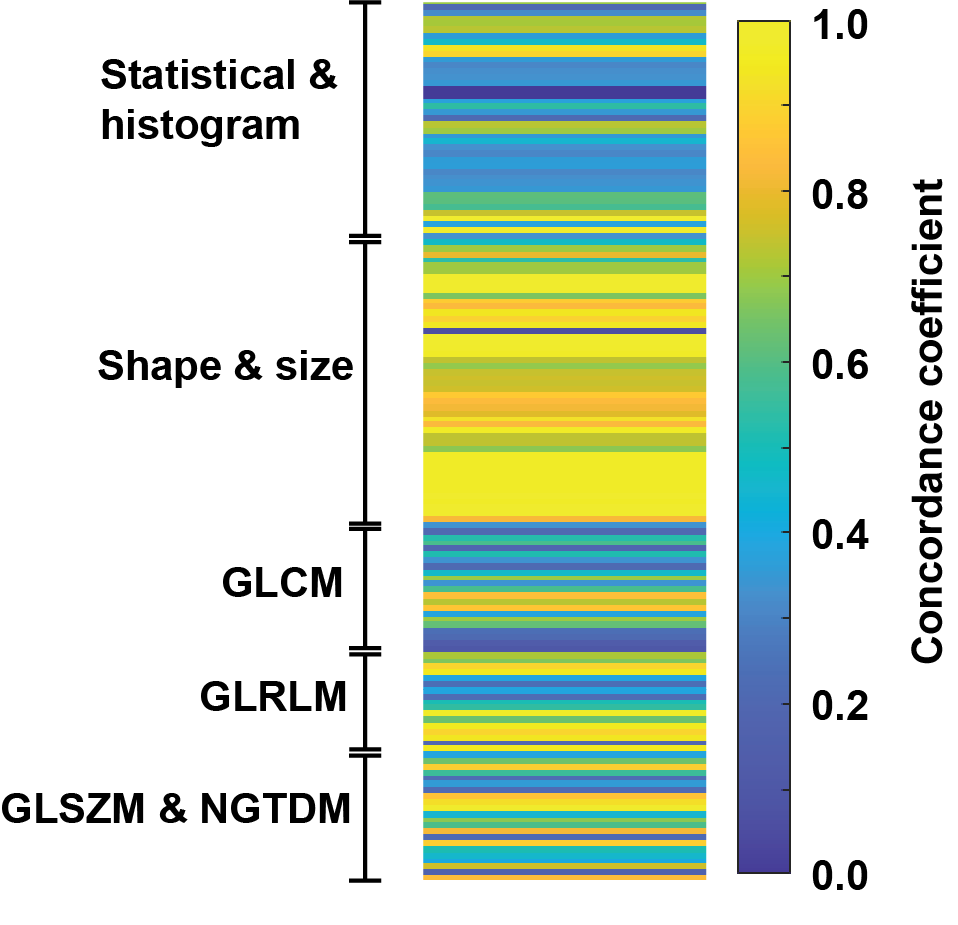

**Supplementary Figure 1.** Feature robustness to tumor segmentation. For all tumors in the training cohort, radiomic features were calculated for the original ROI as segmented by the radiologist, as well as the same ROI shrunk or dilated radially by 1mm. Concordance Coefficient for feature values between the ROIs is presented in the heatmap, with each line representing a radiomic feature, showing differences between feature types, as indicated on the left. GLCM- Grey Level Co-occurrence Matrix, GLRLM- Grey Level Run Length Matrix, GLSZM- Grey Level Size Zone Matrix, NGTDM -Neighboring Gray Tone Difference Matrix.

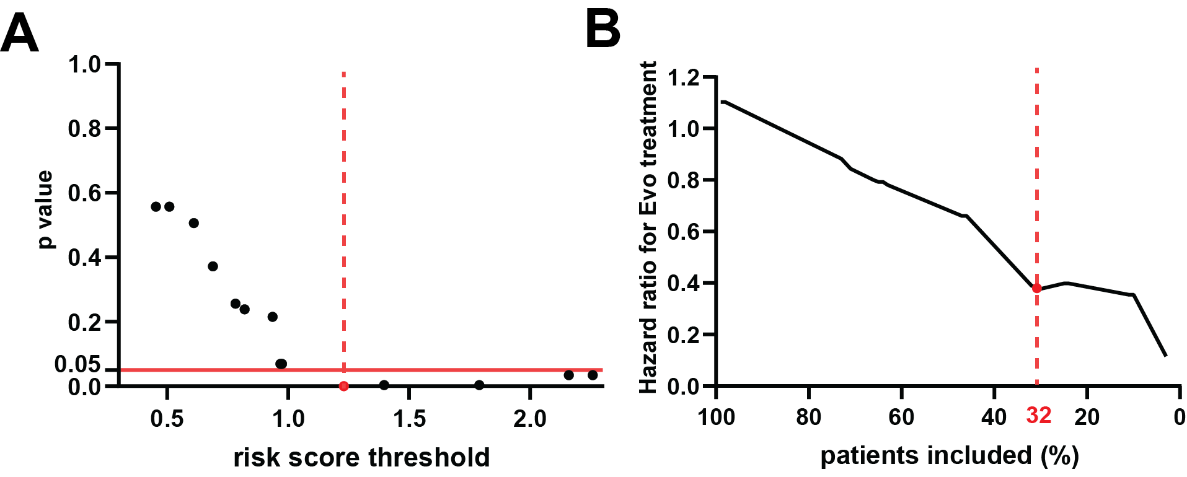

**Supplementary Figure 2. Clinical model performance.** Including only the tumor histology and patient smoking history information in a multivariate Cox model, a risk score threshold was identified for a significantly (p<0.05) different survival in the two treatment groups (A). The graph of corresponding treatment Hazard Ratios (HR) for the considered threshold values (B) shows an increasing benefit of Evo with exclusion of low risk score patients, and 32% of the original training cohort included at the optimal threshold. Solid red line indicates p=0.05 significance level and dashed red line indicates the optimal threshold value and corresponding p and HR.

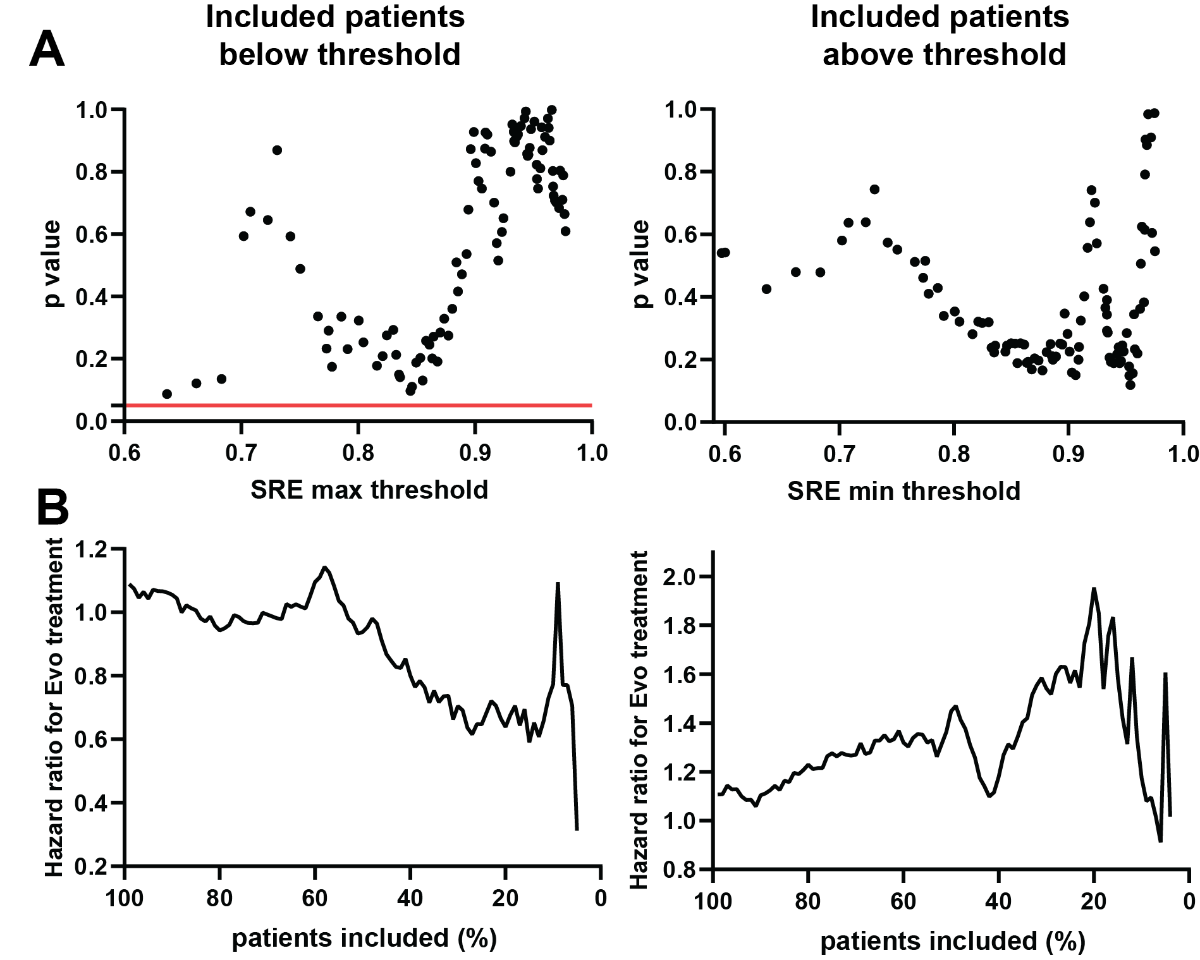

**Supplementary Figure 3. Single feature model performance.** Considering the value of Short Run Emphasis (SRE) radiomic feature as a threshold for inclusion, the graph of p value of survival differences between treatment arms depending on the threshold value is presented in (A), while the corresponding Hazard Ratio plotted against a percentage of patients included at this threshold is shown in (B). The graphs on the left describe the approach where patients with SRE below the threshold are included in the analysis, showing an improved survival of Evo treated patients (HR<1), while the graphs on the right, patients with SRE over the threshold are considered, favoring Dox treatment (HR>1). Solid red line indicates p=0.05 significance level and dashed red line indicates the optimal threshold value and corresponding p and HR.

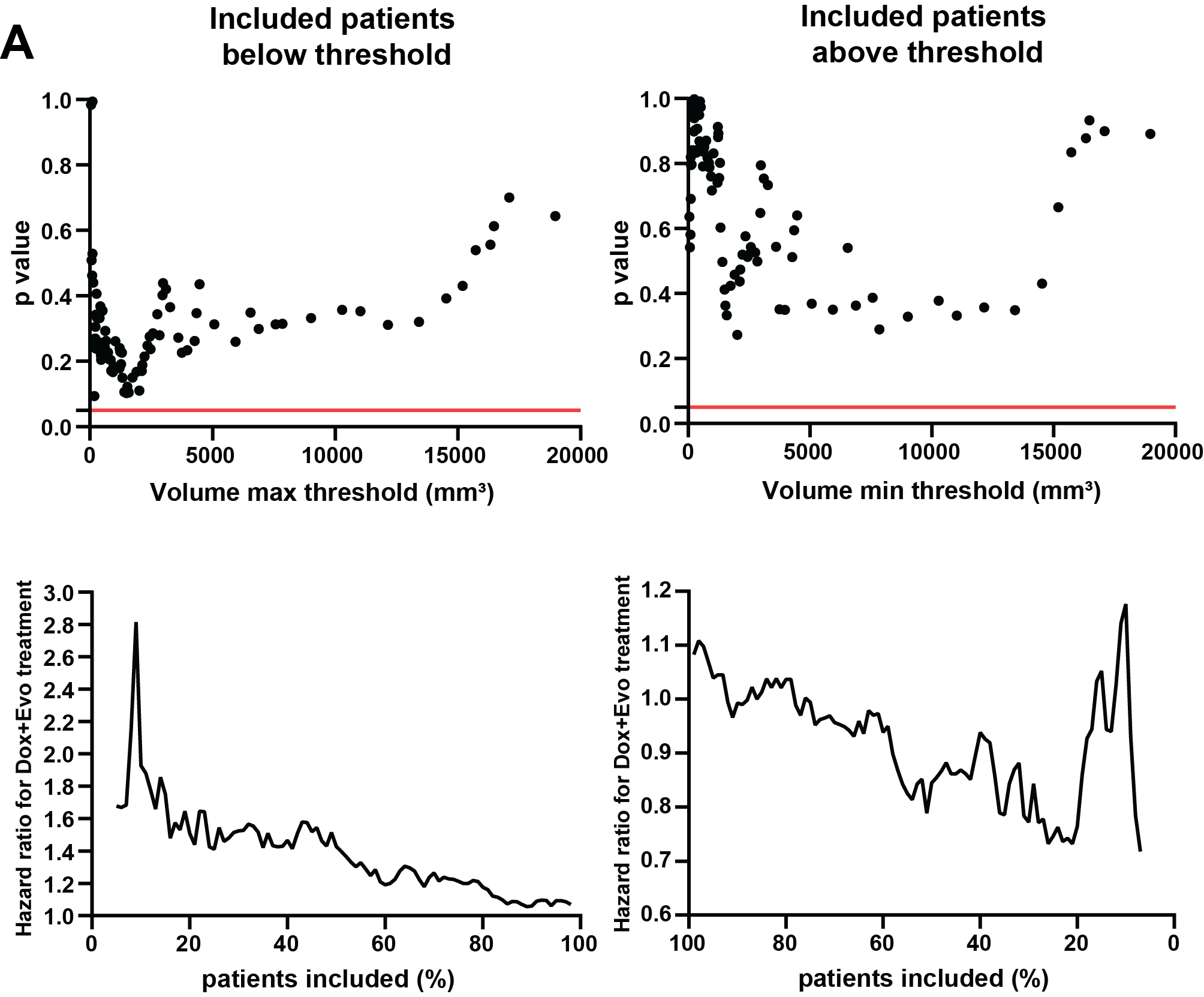

**Supplementary Figure 4. Tumor volume cannot be used for patient selection.** Graphs of p value of survival differences between treatment arms in the treatment cohort depending on the volume threshold value are presented for patient cohort below the maximum volume threshold (A) and above minimum volume threshold (B). Neither of these graphs show points reaching 0.05 value, as indicated by red line. The corresponding Hazard Ratio values for Dox+Evo treatment depending on the percentage of entire training cohort are shown in (C) and (D) for patient inclusion below and above threshold value respectively.

**Supplementary Tables**

| **Feature name** | **Description** |
| --- | --- |
| Age | Age at randomization |
| Sex | Sex |
| Smoking History | History of tobacco smoking, past or present |
| Primary Tum Site | Site of primary tumor |
| Metastatic Sites # | Number of distinct metastatic sites |
| Lung lesions # | Number of segmentable lesions in the lungs |
| Stage | Stage at diagnosis |
| Histology | Histological classification of the primary lesion |
| Tumor Grade | Histological grade of primary tumor |
| ECOG Score | Eastern Cooperative Oncology Group (ECOG) score at diagnosis |
| Prior radiotherapy | Previously treated with radiotherapy (Yes/No) |
| Prior systemic therapy | Treated with other systemic therapy prior to trial (Yes/No) |

**Supplementary Table 1. Description of clinical features.**

**(in Excel document)**

**Supplementary Table 2. Correlation to volume and spatial stability of radiomic features.** The Pearson correlation coefficient and concordance coefficient for spatial stability analysis are shown for all radiomic features.

|  | |  |  |  |
| --- | --- | --- | --- | --- |
|  | |  | **Training** | |
|  | |  | **HR (95% CI)** | **p value** |
| **Histology (vs. Leiomyosarcoma)** | | |  |  |
|  | | Epithelioid | 1.00 (0.228-4.41) | 1.00 |
|  | | Liposarcoma | 1.09 (0.309-3.83) | 0.93 |
|  | | Malignant peripheral nerve sheath tumor | 10.11 (3.31-30.9) | **0.001** |
|  | | Myxofibrosarcoma | 0.665 (0.151-2.92) | 0.59 |
|  | | Pleomorphic rhabdomyosarcoma | 1.50 (0.198-11.4) | 0.70 |
|  | | Pleomorphic sarcoma/ Malignant fibrous histicytoma | 2.29 (1.07-4.89) | **0.03** |
|  | Other | | 1.77 (0.983-3.2) | 0.06 |
| **Smoking history (ever vs. never)** | | | 1.64 (0.95-2.82) | 0.08 |
| **Short Runs Emphasis** | | | 0.023 (0.0018-0.296) | **0.004** |

**Supplementary Table 3. Multivariable Cox model.** The Hazard Ratios (HR) together with 95% Confidence Intervals (CI) and the associated p values (log-rank test) in the multivariable Cox regression model, in the Doxorubicin arm of the training cohort. The model was further applied to identify patients expected to benefit from Doxorubicin monotherapy. Hazard Ratios for categorical variables were calculated against most common category.
